# A data-driven consensus framework for Ct interpretation in real-world multi-assay qPCR diagnostics

**DOI:** 10.64898/2026.06.11.26355491

**Authors:** Jiasi Wang, Jin Chen, Bo Zhao, Guanbin Zhang, Shuang Jian, Tao Deng, Dong Liang

## Abstract

While cycle threshold (Ct) values from quantitative PCR (qPCR) serve as the gold-standard indicators of target abundance, their clinical interpretation is frequently confounded by inherent variability across diverse assay designs, reagents, and instrumentation. In this study, we present a data-driven consensus framework for Ct evaluation that uses large-scale, multi-assay amplification data to establish reference patterns of normal Ct behavior. Based on a total of 41,770 amplification curves collected from four routine diagnostic assays across two PCR platforms, we evaluated machine learning models across three experimental scenarios: within-platform validation, cross-assay generalization, and cross-platform transfer. Extreme gradient boosting (XGBoost) achieved the most accurate and stable predictions under data-sufficient, within-platform conditions with a mean absolute error (MAE) of 0.0419, while pooled multi-assay training improved cross-assay robustness compared with single-assay models. Model performance was further assessed using a deviation-based metric to quantify differences between predicted and instrument-reported Ct values, allowing efficient identification of anomalous amplification curves in large datasets. Notably, direct application across platforms without recalibration led to a substantial decline in performance, with a MAE of 2.62, showing platform-dependent variability. These findings indicate strong stability under within-platform and cross-assay conditions, with scalability contingent upon appropriate cross-platform calibration.

## 1 Introduction

In real-world clinical laboratories, Ct values are not generated under idealized or uniform conditions. Instead, they emerge from heterogeneous combinations of assay designs, reagent systems, amplification chemistries, instruments, and operational environments. As a result, the same biological sample may yield substantially different Ct values across assays or platforms, challenging the assumption that Ct values are intrinsically comparable across assays or platforms. [1,2].

This lack of inherent comparability has important practical consequences. In high-throughput diagnostic workflows, laboratories must routinely interpret Ct values to assess the reliability of results, identify abnormal amplification behaviors, and ensure consistency across batches, assays, and instruments. Existing quality control strategies, such as MIQE-based guidelines [3], technical replicates, and manual curve inspection, provide essential safeguards but are fundamentally limited in scalability and objectivity. These approaches rely heavily on predefined rules or expert judgment and are typically assay- or platform-specific. As testing volumes and assay diversity increase, the complexity of Ct interpretation increasingly exceeds what can be consistently managed through rule-based or manual processes alone.

Recent studies have explored machine learning approaches for analyzing qPCR amplification curves, like automated Ct estimation and curve classification [4]. These methods demonstrate the potential of data-driven modeling [5], but mostly focus on optimizing prediction accuracy for individual assays or controlled datasets. In real-world diagnostics, however, the central challenge is not merely predicting a Ct value, but determining whether a given Ct value is reasonable within the context of historical assay behavior. From this perspective, Ct interpretation can be reframed as a consensus problem: a Ct value is considered reliable if it conforms to the data-driven consensus established by large-scale, previously observed amplification patterns under normal experimental conditions.

In this study, we propose a data-driven consensus framework for Ct interpretation using multi-assay qPCR data. By capturing the implicit consensus of Ct-related amplification behaviors, the framework provides a reference for evaluating new qPCR results and detecting potential anomalies.

## 2 Methods

### 2.1 Data sources, assays, and PCR platforms

#### 2.1.1 Data sources and assays

This study included real-world qPCR amplification data from four diagnostic assays routinely used in clinical molecular testing: hand-foot-mouth disease (HFMD) assay, respiratory multiplex panel (six respiratory pathogens), COVID-19 assay (Maccura reagent kit, SARS-CoV-2 Fluorescent PCR Kit, Maccura Biotechnology Co., Ltd, Chengdu, China), and COVID-19 assay (Liferiver reagent kit, Shanghai ZJ Bio-Tech Co., Ltd, Shanghai, China). The Maccura COVID-19 and Liferiver COVID-19 assays represent SARS-CoV-2 detection based on two distinct commercial reagent kits, each employing independent primer–probe designs and reaction chemistries. All four assays were analyzed using a unified post-run computational workflow to assess model robustness across assays with multiple biochemical characteristics.

#### 2.1.2 PCR instruments and platforms

In this study, qPCR amplification data were generated from two real-time PCR instruments: the SLAN-96S (Shanghai Hongshi Medical Technology Co., Ltd, Shanghai, China) and the Gentier 96R (Xi’an Tianlong Science and Technology Co., Ltd, Xi’an, China). The SLAN-96S platform was used for all four diagnostic assays (HFMD, respiratory multiplex, Maccura COVID-19, and Liferiver COVID-19), whereas the Gentier 96R platform was used exclusively for an independent Maccura COVID-19 dataset. This configuration enabled both multi-assay evaluation within a single platform and cross-platform generalization analysis, allowing systematic assessment of instrument-induced domain shifts. To facilitate robust computational modeling, samples lacking instrument-reported Ct values were assigned a terminal value of 40 cycles to represent non-amplification, a procedure consistent with established diagnostic reporting conventions.

To evaluate whether model-derived Ct values are consistent with instrument-reported results in real-world diagnostic settings, we introduce a Ct prediction deviation metric:

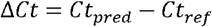

Where Ct_pred denotes the Ct value predicted by the model, and Ct_ref denotes the reference Ct value reported by the qPCR instrument. This ΔCt represents the deviation between model-estimated and instrument-generated Ct values and is distinct from the conventional ΔCt used in expression normalization workflows. The absolute deviation |ΔCt| is used to quantify the magnitude of disagreement.

### 2.2 Feature extraction and normalization

Each amplification curve was represented by 45 extracted features summarizing amplification dynamics and signal characteristics, which were used for model development. To reduce scale-related bias across assays and platforms, feature values were subjected to a unified normalization procedure. Normalization parameters were derived exclusively from training data and applied to all test datasets to prevent information leakage.

#### 2.3 Experimental design and datasets

Three complementary experimental settings were designed to evaluate model performance under realistic diagnostic constraints.

First, to assess model behavior under data-sufficient conditions and to compare different learning strategies, a large-scale dataset from the Maccura COVID-19 assay on the SLAN-96S platform was used. A total of 26,820 samples were randomly divided into a training set (20,116 samples) and an independent test set (6,704 samples, ∼25% of total samples). XGBoost, recurrent neural networks (RNN), and multilayer perceptron regressors (MLPRegressor) were trained using identical feature representations and evaluated on the same test set. To assess robustness to random initialization and data shuffling, each model was trained and evaluated using three different random seeds (1, 4, and 42). Performance metrics were reported for each run.

Second, based on XGBoost, a pooled model was trained using combined data from all four diagnostic assays generated on the SLAN-96S platform to examine cross-assay generalization. The pooled evaluation included independent test sets from four diagnostic assays: Respiratory Multiplex panel (10,896 samples), Maccura COVID-19 (6,704 samples), Liferiver COVID-19 (1,100 samples), and HFMD (90 samples). In parallel, a single-assay model was trained only on the Maccura COVID-19 dataset from the SLAN-96S platform. This model was evaluated on its in-domain test set (Maccura COVID-19, 6,704 samples) and on external test sets from the other three diagnostic assays (Respiratory Multiplex, Liferiver COVID-19, and HFMD) to assess cross-assay transferability under assay-mismatch conditions. Both pooled and single-assay models were evaluated on independent test sets from each diagnostic assay to compare performance stability and generalization capability across multiple assays.

Third, to investigate cross-platform transferability, the model trained on the SLAN-96S platform with the Maccura COVID-19 dataset was directly applied to another independent Maccura COVID-19 dataset generated on the Gentier 96R platform (2,864 samples), without any platform-specific recalibration. This setting simulated real-world deployment scenarios in which trained models are transferred across PCR instruments.

### 2.4 Performance evaluation and implementation

Model performance was quantified using mean absolute error (MAE) and root mean squared error (RMSE). In addition, the proportions of samples with the absolute deviation |ΔCt| (0.5, 1, and 3 cycles) were reported to provide clinically interpretable measures of prediction reliability.

All data processing, model training, and evaluation procedures were implemented in Python (version 3.12) using standard scientific computing libraries.

## 3 Results

Evaluation across three complementary experimental scenarios revealed that Ct prediction stability is fundamentally governed by the interplay between data diversity and platform-specific signal characteristics. Across three complementary evaluation settings, model performance consistently reflected the impact of data diversity and platform variability on Ct prediction stability. Under data-sufficient, within-platform validation conditions, XGBoost achieved the most accurate and stable predictions, with an MAE of 0.0419 across different random seeds. The proportion of samples with |ΔCt| > 0.5 was only 1.48%, and deviations greater than 1 cycle were rare (0.34%). In contrast, MLP showed moderate degradation (MAE 0.17–0.26), while RNN exhibited substantial instability across seeds. These findings indicate that tree-based ensemble learning provided more robust modeling of amplification curve features compared with neural network architectures under identical feature representations [6] (Figure 1).

**Figure 1.**
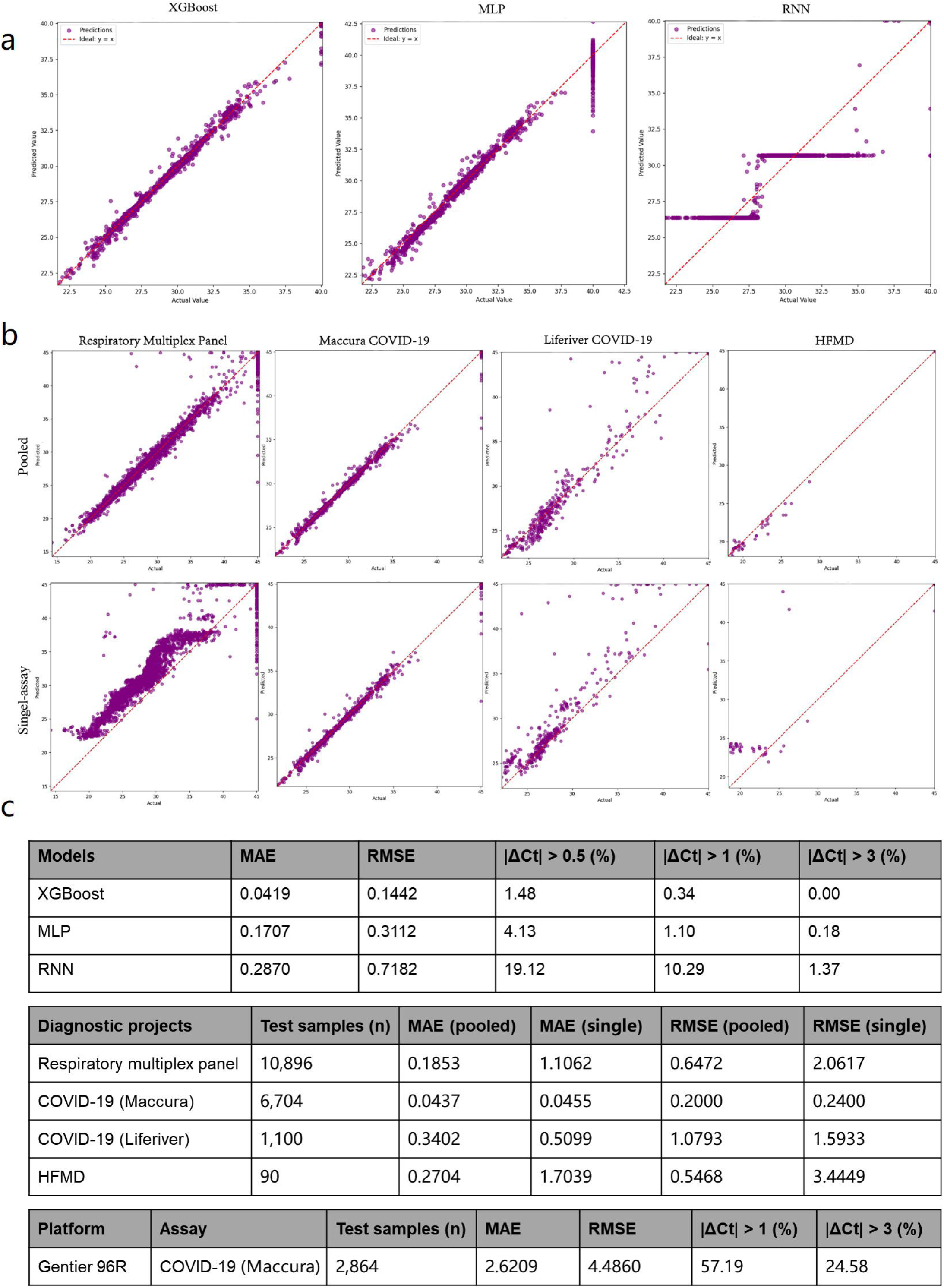
Model performance under within-platform, cross-assay, and cross-platform evaluation settings. (a) Predicted versus instrument-reported Ct values for XGBoost, MLP, and RNN under data-sufficient, within-platform validation (Maccura COVID-19, SLAN-96S). Results are shown for one representative random seed among three independent runs. The red dashed line indicates the identity line (y = x). (b) Cross-assay generalization of pooled and single-assay XGBoost models across Respiratory Multiplex Panel, Maccura COVID-19, Liferiver COVID-19, and HFMD assays (SLAN-96S platform). (c) Quantitative performance summary, including MAE, RMSE, and proportions of samples exceeding absolute deviation thresholds (|ΔCt| > 0.5, 1, and 3 cycles), as well as cross-platform transfer results on the Gentier 96R dataset without recalibration.

Under the cross-assay generalization setting on the same platform, performance differences emerged between the pooled and single-assay XGBoost models when evaluated across different diagnostic assays. On the in-domain Maccura COVID-19 test set, both models achieved comparable prediction accuracy, with similar MAE and comparable |ΔCt| distributions, indicating that the single-assay model retained strong fitting capacity within its training assay. However, when applied to out-of-domain assays (Respiratory Multiplex, Liferiver COVID-19, and HFMD), the single-assay model exhibited increased error dispersion. In contrast, the pooled model maintained relatively consistent error distributions across all four assays. Although minor performance fluctuations were observed, the frequency of large-deviation samples was consistently lower than that of the single-assay model under assay-mismatch conditions. This stability indicates that exposure to multi-assay amplification patterns during training mitigated overfitting to assay-specific signal characteristics and improved cross-assay robustness [7].

Under the cross-platform transfer setting, performance degradation became pronounced when the single-assay model trained on SLAN-96S Maccura COVID-19 data was directly applied to the Gentier 96R Maccura COVID-19 dataset without recalibration. The MAE increased to 2.62 cycles, and 24.58% of samples exhibited |ΔCt| > 3 cycles. This substantial shift suggests that amplification behavior learned from one PCR platform does not directly generalize to another. Further analysis revealed that the degradation was primarily driven by a systematic offset—likely due to platform-specific optical sensitivities—rather than unpredictable stochastic noise. Collectively, these results demonstrate a clear hierarchy of stability: strong agreement under within-platform, data-sufficient validation; improved cross-assay robustness through pooled training; and a marked decline in performance under cross-platform transfer that necessitates instrument-specific calibration or fine-tuning. The findings indicate that both assay diversity and platform-specific signal characteristics strongly condition Ct prediction stability.

## 4 Discussion

Unlike prior studies that focus on optimizing Ct for individual amplification curves, our approach treats Ct as a population-level statistical quantity learned from large-scale, real-world amplification data. Rather than fitting a deterministic function to each curve, the model captures the implicit consensus of expert interpretation and system-specific experimental variability. This enables accurate Ct prediction and the identification of abnormal amplification behaviors that deviate from historical distributions [8,9], addressing limitations of traditional rule-based and curve-fitting approaches in practical clinical settings.

Within a single assay and platform, XGBoost captured amplification dynamics accurately and consistently, outperforming neural network architectures in both prediction stability and error magnitude. Multi-assay training further improved robustness, reducing large deviations when applied to out-of-domain assays and mitigating overfitting to assay-specific signal characteristics. Exposure to diverse amplification patterns during training enhances cross-assay generalization while maintaining in-domain prediction accuracy.

An important feature of our framework is the |ΔCt| metric, which quantifies deviations between model-predicted and instrument-reported Ct values. This measure provides a practical, threshold-based approach to flag anomalous amplification curves [10]. In a retrospective expert review of representative cases with |ΔCt| > 3, we observed that these large deviations were frequently associated with atypical curve morphologies, including baseline drift, non-specific amplification, or reduced signal-to-noise ratios. This provides preliminary evidence supporting the utility of the framework as an objective “second opinion” for high-throughput screening in large clinical datasets. Importantly, the goal of the machine learning model is not to replace laboratory personnel or manual quality control, but to complement existing workflows. By combining model predictions with expert review, laboratories can prioritize samples that require attention, reduce variability arising from subjective judgment, and improve overall diagnostic efficiency, particularly in high-volume testing environments.

Still, the cross-platform transfer remains a challenge. Substantial prediction errors were observed when models trained on one instrument were applied to data from another without recalibration, reflecting systematic differences in amplification behavior across platforms rather than random noise. Platform-specific calibration or adaptive modeling is therefore necessary for reliable deployment in different diagnostic settings. This limitation will be addressed in our future work.

In summary, our findings establish a clear hierarchy of predictive stability, defining the operational boundaries for data-driven Ct interpretation across varying levels of assay and platform heterogeneity. Strong agreement is observed under within-platform, data-sufficient conditions, and pooling data from multiple assays improves cross-assay robustness. Nevertheless, it’s important to apply careful adaptation for cross-platform applications. By providing scalable, objective references for normal Ct behavior and a systematic approach for anomaly detection, the data-driven consensus modeling can optimize workflow efficiency, support quality control, and enhance confidence in real-world qPCR diagnostics.

## Data Availability

All data produced in the present study are available upon reasonable request to the authors

http://labeasy.cn/

## Ethics Statement

This study focused on the methodological development and validation of machine learning models for Ct value determination using qPCR amplification curve data. All datasets were retrospectively collected and fully anonymized prior to analysis, with no access to personal identifiable information or clinical records.

## Data and Code Availability

The datasets and source code used in this study are not publicly available due to commercial confidentiality and ongoing intellectual property protection. However, relevant materials may be made available from the corresponding author upon reasonable request and with permission from the data-owning institution.

## Patents

This study is based on our previously granted Chinese patent.

Ping Tang, Bo Zhao, Dong Liang, Zhi Yan, Tao Jiang. A Method and System for Ct Value Detection in PCR. ZL 202411108830.6, China, granted 2025-02-11.

